# Disparities in seizure outcomes revealed by large language models

**DOI:** 10.1101/2023.09.20.23295842

**Authors:** Kevin Xie, William K.S. Ojemann, Ryan S. Gallagher, Alfredo Lucas, Chloé E. Hill, Roy H. Hamilton, Kevin B. Johnson, Dan Roth, Brian Litt, Colin A. Ellis

## Abstract

**Objective:** Large-language models (LLMs) in healthcare have the potential to propagate existing biases or introduce new ones. For people with epilepsy, social determinants of health are associated with disparities in access to care, but their impact on seizure outcomes among those with access to specialty care remains unclear. Here we (1) evaluated our validated, epilepsy-specific LLM for intrinsic bias, and (2) used LLM-extracted seizure outcomes to test the hypothesis that different demographic groups have different seizure outcomes.

**Methods:** First, we tested our LLM for intrinsic bias in the form of differential performance in demographic groups by race, ethnicity, sex, income, and health insurance in manually annotated notes. Next, we used LLM-classified seizure freedom at each office visit to test for outcome disparities in the same demographic groups, using univariable and multivariable analyses.

**Results:** We analyzed 84,675 clinic visits from 25,612 patients seen at our epilepsy center 2005-2022. We found no differences in the accuracy, or positive or negative class balance of outcome classifications across demographic groups. Multivariable analysis indicated worse seizure outcomes for female patients (OR 1.33, p = 3x10^-8^), those with public insurance (OR 1.53, p = 2x10^-13^), and those from lower-income zip codes (OR ≥ 1.22, p ≤ 6.6x10^-3^). Black patients had worse outcomes than White patients in univariable but not multivariable analysis (OR 1.03, p = 0.66).

**Significance:** We found no evidence that our LLM was intrinsically biased against any demographic group. Seizure freedom extracted by LLM revealed disparities in seizure outcomes across several demographic groups. These findings highlight the critical need to reduce disparities in the care of people with epilepsy.

**Key Points:**

- We used large language models (LLMs) and natural language processing to extract seizure outcomes from clinical note text.
- We found no evidence of intrinsic bias in the LLM algorithm, in that it performed similarly across all demographic groups.
- Using LLM-extracted seizure outcomes, female sex, public insurance, and lower income zip- codes were associated with higher likelihood of seizures at each visit.
- Black race was associated with higher likelihood of seizures in univariable but not multivariable analysis.
- These findings highlight the critical need to reduce disparities in the care of people with epilepsy.

## Introduction

In the US healthcare system, minoritized and marginalized groups experience profound disparities in access to care and health outcomes.^1^ In neurology, for example, non-White patients have less access to neurologic care than White patients and worse health outcomes across many neurologic conditions and subspecialties.^2, 3^ For people with epilepsy, demographic factors play a role in disease prevalence, access to specialty care, surgical evaluation, and in-hospital mortality.^4–7^ However, most studies of disparities in epilepsy have focused on access to care.^7^ Relatively little is known about how demographic factors impact critical outcomes, such as treatment response and seizure freedom.

Just as disparities exist in healthcare, there are well-founded concerns that artificial intelligence tools used to deliver or evaluate healthcare will propagate existing biases or introduce new ones.^8–10^ For example, a widely used commercial risk prediction tool to guide allocation of care resources was found to underestimate the severity of illness for Black patients relative to White patients, exacerbating a disparity in access to care.^11^ One form of artificial intelligence, Natural Language Processing (NLP), uses machines to read and understand human language, which enables rapid, large-scale extraction and analysis of unstructured clinical data from electronic health records. NLP is vulnerable to perpetuated bias because biases present in unstructured note text and in healthcare delivery more broadly will be incorporated into the NLP algorithm. For example, a recent algorithm for identifying opioid misuse using clinical notes was found to have bias in Black compared to White individuals.^12, 13^ Recent developments in ethical AI have emphasized the importance of ensuring equality in model performance between protected groups.^8, 10, 11, 14, 15^

We have recently developed and validated an NLP algorithm to extract epilepsy outcome measures from the unstructured text of clinical notes with accuracy similar to human readers.^16–18^ In this study, we first tested our algorithm for bias, in the form of differential performance in different demographic groups. An unbiased NLP algorithm was a prerequisite to our second objective, which was to use the algorithm to test the hypothesis that different demographic groups have different likelihoods of seizure freedom at our academic US healthcare center.

## Materials and Methods

This research was approved by the Institutional Review Board of the University of Pennsylvania with a Waiver of Informed Consent.

### Data Collection

Our source dataset consisted of electronic health records from patients who had seen an epileptologist at the University of Pennsylvania Health System between the years of 2005 and 2022. From these records, we obtained clinical notes written by their epileptologist(s) with full text and date; medication prescriptions; and demographic information including race, sex, zip code, age, and insurance provider at each visit. We excluded patients with missing demographic data.

### Outcomes: seizure classification by NLP

We recently developed and validated an NLP algorithm that classifies clinic notes as seizure-free or having recent seizures.^16, 17^ Briefly, we used Clinical_BERT,^19^ a publicly-available transformer language model from Google AI, on 700 manually annotated epileptologist notes. We defined a “seizure free” visit as one where the patient did not have seizures since their last visit, or within the past year, whichever was more recent. Model predictions were repeated five times using different seeds, and final classification of each note was determined by plurality voting of the five outputs.

### Exposures: Demographic Variables

Race, ethnicity, and sex were self-reported by patients at the time of initial registration with our health system, while zip code and insurance were entered and verified by clinic registration staff. Category options were determined by the Epic electronic health record software (Epic, Madison WI). Race category options were as follows, listed alphabetically: American Indian or Alaskan Native; Asian; Black or African American; East Indian; Native Hawaiian or Other Pacific Islander; Some Other Race; White.

Only Asian, Black, and White had sufficient samples for analysis, and the other categories were combined into a category of “Other Races.” Ethnicity categories were Hispanic Latino or Not Hispanic Latino. Sex categories were female or male. Gender identities, including trans and non-binary genders, were not recorded.

Zip codes were linked to median household income, in inflation-adjusted 2020 dollars, using publicly available data from the US Census (data.census.gov). We grouped zip codes into four categories according to median household income: (1) less than $50,000; (2) $50,000 to less than $75,000; (3) $75,000 to less than $100,000; (4) $100,000 or more. Insurance type was classified as private insurance, Medicare, or Medicaid. For purposes of analysis we combined Medicare and Medicaid into a single category of public insurance. Age was grouped into the following categories: 18-39, 40-64, and 65 or older.

### Assessing Model Bias

These analyses used 192 manually-annotated notes from the validation dataset from our previous study.^16^ Human annotations were performed in triplicate by independent readers who were unaware of the identities or demographics of the patients. Here we sought to determine whether our NLP models performed differently in different demographic groups using several measures. First, we calculated the accuracy of the model classifications (seizure free vs. recent seizure), where accuracy was the number of correct classifications divided by all classifications (**Supplemental Methods**). Second, we calculated the positive class balance (PCB) and negative class balance (NCB) of each demographic group.^15, 20^ These methods use the probability values that accompany each model prediction (for example, the model may predict that a note describes recent seizures with probability of 0.92). PCB is the average predicted probability of recent seizures in patients who did have recent seizures, with an expected value near 1. NCB is the average predicted probability of recent seizures in patients who did not have recent seizures, with an expected value near 0. If the NLP model made predictions with perfect confidence, the PCB would be the complement of the false negative rate (FNR = 1 - PCB) and the NCB would be the equivalent of the false positive rate (FPR = NCB).

### Statistical Analysis

For the analysis of model bias, we tested for differences in accuracy, PCB, and NCB between demographic groups using Fisher’s exact tests (accuracy) or two-tailed permutation tests with 10,000 iterations (class balances) for the binary demographic variables (sex, ethnicity, and insurance), and using two-sided Kolmogorov-Smirnov (KS) tests against the null hypothesis of a uniform distribution for other categorical variables (race, income, and age).

For testing associations between demographics and seizure freedom classification, we performed a series of logistic mixed effects regression models. In each model the outcome was the seizure freedom classification of each visit; exposure was the demographic variable; and patient was the clustering variable (random effect) to account for intra-individual correlation across repeated measures. We included the time (in months) since last visit as a covariate in each model, to account for the fact that patients with frequent seizures are likely to be seen more often than seizure-free patients.^18^ Additionally, we included the number of ASMs a patient was prescribed at the time of a visit as a variable of interest to act as a positive control, as we expected to find an association between seizure freedom likelihood and number of ASMs.

First we tested each demographic variable in a separate mixed effects model, representing “univariable” analyses, adjusting only for time since last visit, and patient-specific random effect. Second, we combined all demographic variables in a single mixed-effects model, representing a multivariable analysis to assess the competing effects of demographic variables.

We adjusted p-values for multiple comparisons in both the regression and bias univariable analyses using the Benjamini-Hochberg false discovery rate method with an ⎕of 0.05.^21^

We included additional methodological details within our supplement. All analyses were performed with Python and used the following packages: transformers, statsmodels, numpy, pandas, scipy, forestplot, and pymer4. Our NLP models are available on the Hugging Face hub at https://huggingface.co/CNT-UPenn, and our code is available on GitHub at https://github.com/penn-cnt/NLP_Disparities_in_Seizure_Freedom. We do not make our data available to protect patient privacy.

## Results

### Cohort

Demographic information was available for 25,612 patients and is summarized in **Table 1**. Of the 84,675 visits from which seizure freedom could be determined, 22,038 (26%) were classified as seizure free and 48,327 (57%) were classified as having recent seizures. A total of 3,265 patients were excluded due to missing or incomplete demographics and 14,310 visits were excluded due to unclassifiable seizure freedom. To visualize the spatial distribution of our patient cohort in our local tri-state area, we have also made interactive maps of our patients and some of their demographic variables, by zipcode **(Supplemental Results)**.

**Table 1:** Summary of patient-reported demographic information of our patient cohort.

| Demographic | N (%) of 25,612 Patients |
| --- | --- |
| Race |  |
| American Indian or Alaskan Native | 67 (0.3%) |
| Asian | 741 (2.9%) |
| Black or African American | 8,308 (32.4%) |
| East Indian | 6 (0.0%) |
| Native Hawaiian or Other Pacific Islander | 15 (0.1%) |
| Some Other Race | 990 (3.9%) |
| White | 13,916 (54.3%) |
| Ethnicity |  |
| Hispanic Latino | 826 (3.2%) |
| Not Hispanic or Latino | 24,313 (94.9%) |
| Sex |  |
| Male | 11,078 (43.3%) |
| Female | 14,534 (56.7%) |
| Age at Latest Visit |  |
| 18-39 | 7,785 (30.4%) |
| 40-64 | 10,081 (39.4%) |
| 65 or older | 7,746 (30.2%) |
| Insurance |  |
| Public | 14,830 (57.9%) |
| Private | 9,227 (36.0%) |
| Median Income per Zip Code |  |
| Less than \$50,000 | 6,960 (27.2%) |
| \$50,000 to <\$75,000 | 5,646 (22.0%) |
| \$75,000 to <\$100,000 | 6,452 (25.2%) |
| Over \$100,000 | 6,384 (24.9%) |

### No evidence of model bias

We found no evidence of model bias with respect to the demographic variables analyzed here (**Figure 1** and **Supplementary Table 1**). Specifically, the accuracy of the NLP model did not differ according to sex (Fisher’s Exact adj. p = 0.96), race (KS adj. p = 0.96), ethnicity (Fisher’s Exact adj. p = 1.00), insurance (Fisher’s Exact adj. p = 0.86), income (KS adj. p = 0.86), or age (KS adj. p = 0.86).

**Figure 1:**
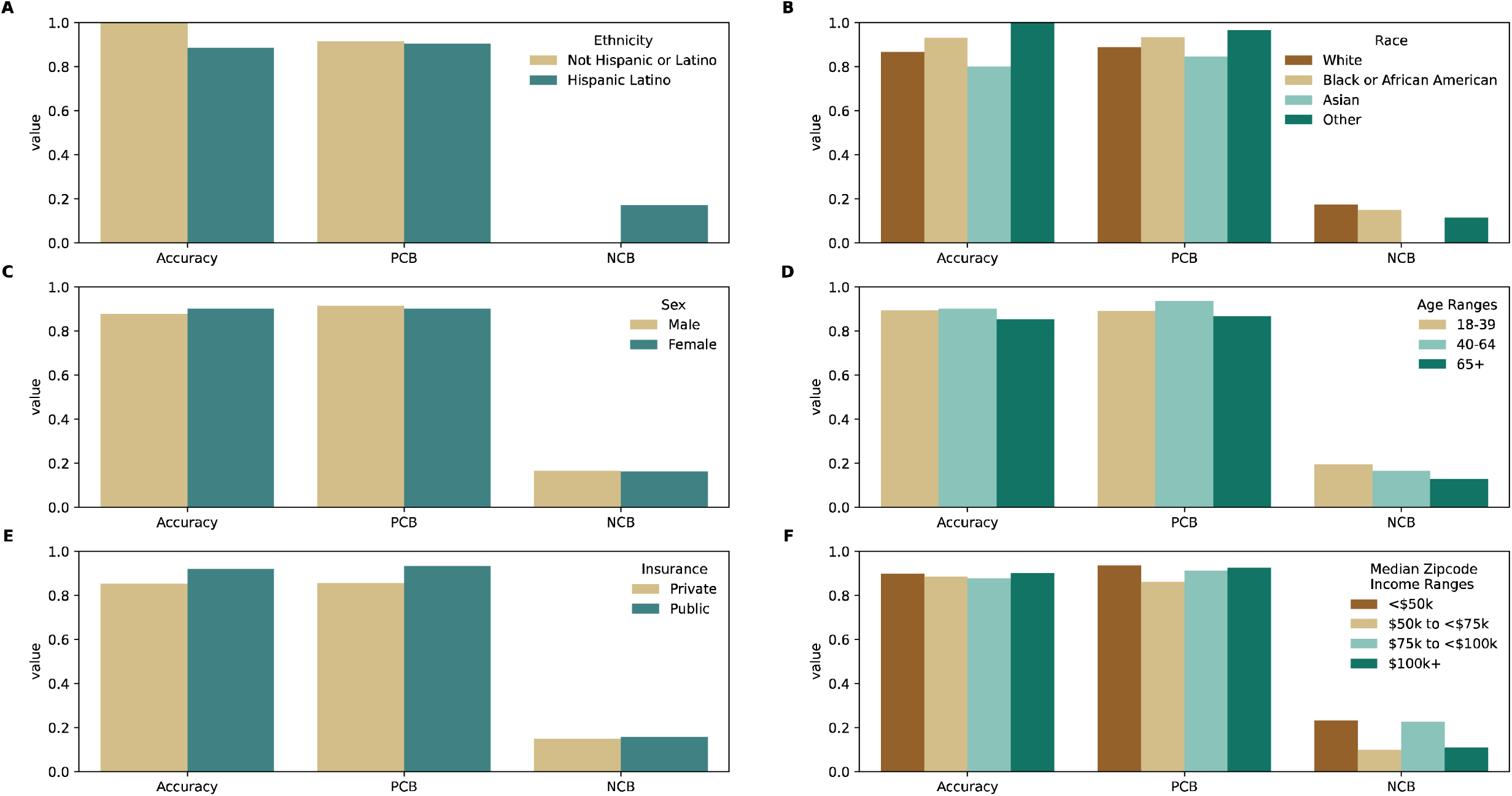
Assessing Model Bias. Model accuracy, positive class balance (PCB), and negative class balance (NCB) stratified by demographic variables. (A) Ethnicity, (B) Sex, (C) Insurance, (D) Race, (E) Age, (F) Income All statistical comparisons between groups were non-significant (see text).

Furthermore, PCB and NCB did not differ according to sex (permutation test adj. p = 0.86 and 0.96, respectively), race (KS adj. p = 0.96 and 0.86), ethnicity (permutation test adj. p = 0.96 and 0.86), insurance (permutation test adj. p = 0.86 and 0.86), income (KS adj. p = 0.96 and 0.86), or age (KS adj. p = 0.96 and 0.96).

### Disparities in seizure freedom classification

Disparities in the likelihood of seizure-free visits were present for each demographic variable we analyzed, with the minoritized and marginalized groups having worse outcomes than privileged groups (**Figure 2**). Specially, female patients (OR 1.35, 95% CI 1.21-1.50, adj. p = 7.9x10^-8^), Black patients (OR 1.40, 95% CI 1.24-1.58, adj. p = 1.4x10^-7^), patients with public insurance (OR 1.48, 95% CI 1.33-1.64, adj. p = 4.6x10^-12^), and patients living in zip codes with less than $100,000 median income (OR ≥ 1.17, adj. p ≤ 0.037), were more likely to have recent seizures than patients from privileged groups. Older patients were more likely to be seizure free than younger patients. Greater numbers of prescribed ASMs were associated with lower likelihood of seizure freedom, as expected.

**Figure 2:**
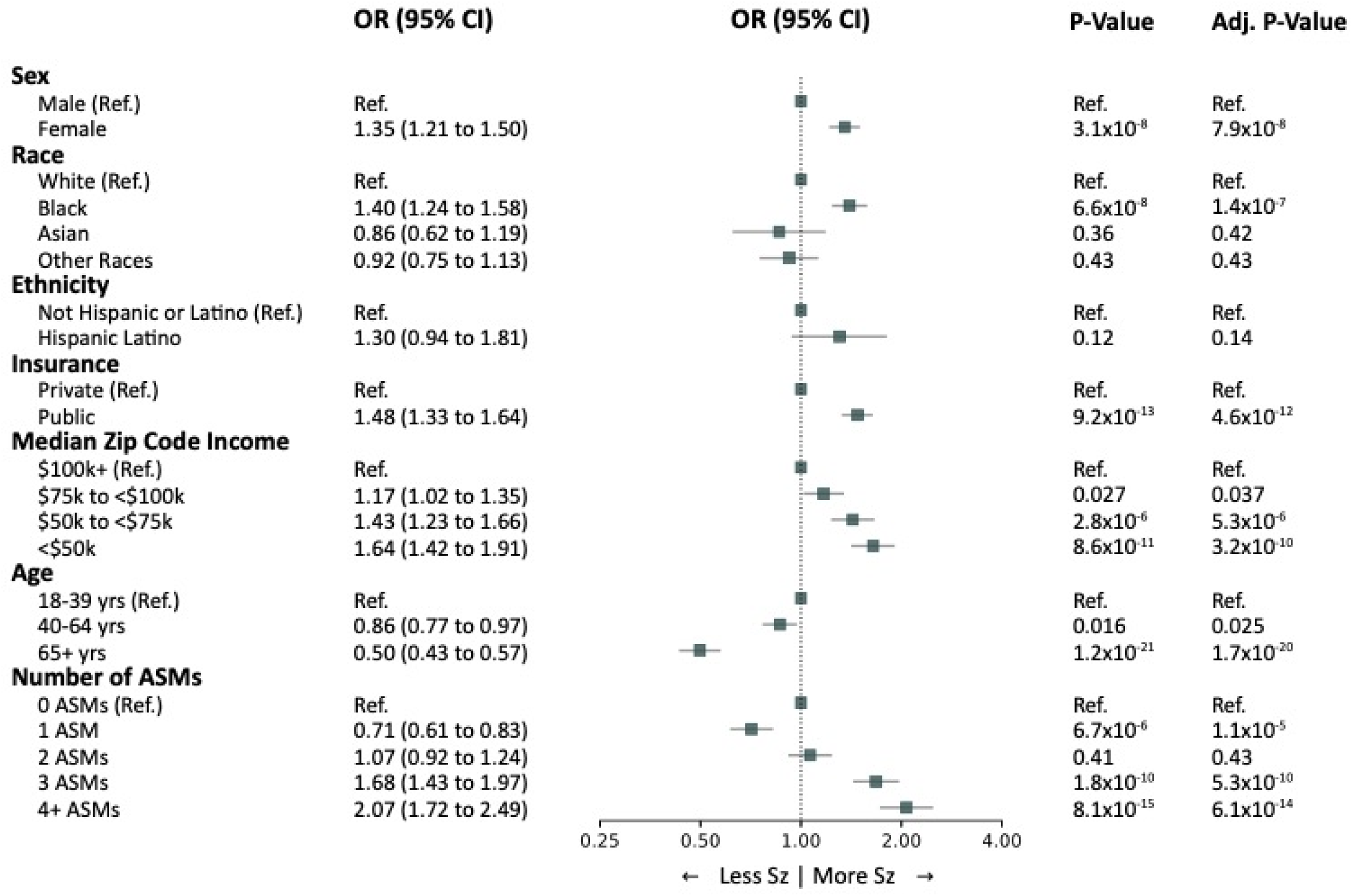
Disparities in seizure freedom likelihood. Forest plot with univariable estimates of odds ratios (OR) of recent seizures at each office visit. ORs were estimated from logistic mixed model regression at the visit level, grouping visits by patients, and controlling for time since last visit. “Other Races” included “American Indian or Alaskan Native”, “East Indian”, “Native Hawaiian or Other Pacific Islander”, and “Some Other Race.” Adjusted p-values were calculated using the Benjamini-Hochberg false discovery rate correction with an ⎕0.05.

Some but not all of these disparities persisted in the multivariable regression analysis (**Table 2**). Black patients no longer had significantly higher odds of seizures, and patients of Other Races had significantly lower odds of seizures than White patients, indicating some overlap between the effects of race and other demographic variables.

**Table 2:** Multivariable Analysis of Demographic Variables and Seizure Freedom.

| Variable | Estimate | OR | 95% CI | P-value |
| --- | --- | --- | --- | --- |
| Sex<br>(Ref. Men) | 0.28 | 1.33 | 1.20 - 1.46 | $3.2 \times 10^{-8}$ |
| Race<br>(Ref. White) |  |  |  |  |
| Black | 0.03 | 1.03 | 0.89 - 1.19 | 0.66 |
| Asian | -0.21 | 0.81 | 0.60 - 1.09 | 0.17 |
| Other Races | -0.22 | 0.80 | 0.65 - 0.98 | 0.035 |
| Ethnicity<br>(Ref. Not Hispanic or Latino) | 0.15 | 1.16 | 0.83 - 1.61 | 0.38 |
| Insurance Type<br>(Ref. Private) | 0.42 | 1.53 | 1.37 - 1.71 | $1.5 \times 10^{-13}$ |
| Median Zip Code Income*<br>(Ref. \$100k+) | | | | |
| \$75k to <\$100k | 0.11 | 1.11 | 0.97 - 1.27 | 0.11 |
| \$50k to <\$75k | 0.20 | 1.22 | 1.06 - 1.41 | $6.6 \times 10^{-3}$ |
| <\$50k | 0.28 | 1.33 | 1.11 - 1.58 | $1.4 \times 10^{-3}$ |
| Age*<br>(Ref. 18-39 yrs) |  |  |  |  |
| 40-64 yrs | -0.23 | 0.80 | 0.71 - 0.89 | $7.1 \times 10^{-5}$ |
| 65+ yrs | -0.84 | 0.43 | 0.37 - 0.50 | $6.4 \times 10^{-31}$ |
| Number of ASMs Taken (Ref. 0 ASMs) |  |  |  |  |
| 1 ASM | -0.33 | 0.72 | 0.62 - 0.83 | $1.0 \times 10^{-5}$ |
| 2 ASMs | 0.05 | 1.05 | 0.91 - 1.23 | 0.49 |
| 3 ASMs | 0.49 | 1.64 | 1.39 - 1.92 | $1.3 \times 10^{-9}$ |
| 4 ASMs | 0.69 | 1.99 | 1.65 - 2.39 | $2.3 \times 10^{-13}$ |
| Months Since Last Visit | -0.02 | 0.98 | 0.98 - 0.99 | $1.0 \times 10^{-19}$ |

## Discussion

In this study, using NLP-derived seizure freedom data from over 25,000 patients with epilepsy across nearly 85,000 clinic visits, we found disparities in the likelihood of seizure freedom according to demographic factors. Female sex, Black race, public insurance, and lower income zip-codes were all associated with higher likelihood of seizures at each clinic visit, controlling for the time between visits. The disparities by sex, insurance, and income persisted after controlling for other demographic factors.

We did not find evidence of bias in our algorithm, in that the models performed similarly across all demographic groups. This work aligns with the American Medical Association’s policy recommendations that emphasized the development of health care AI with a mind to identify biases and prevent exacerbation of health disparities.^9, 22^ Our finding is particularly important because other pretrained transformer models have exhibited bias in several contexts.^23, 24^ The observation that our model was unbiased suggests that disparities in seizure freedom likelihood were not influenced by model failures.

Our findings indicate that seizure freedom likelihood is influenced by demographic factors. Prior studies of disparities in the care of patients with epilepsy have focused mainly on access to specialty care,^4, 6, 7, 25^ with fewer studies examining disparities in epilepsy outcomes. Black patients are less likely to undergo epilepsy surgery than White or Hispanic patients,^26^ but one study of outcomes after temporal lobe epilepsy surgery in 252 patients found no differences according to race or socioeconomic status.^27^ Although socioeconomic factors are associated with differences in ASM adherence,^28–32^ the downstream impact on epilepsy outcomes have not been reported. A study of adults with epilepsy in the Swedish population registry found that lower income and education were associated with more hospitalizations and less access to neurologists.^33^ A study of 1,038 pediatric patients found that Hispanic/Latino ethnicity and lower income zip codes were associated with seizures that worsened over time.^34^ Our findings add to the growing awareness that health outcomes of people with epilepsy vary with demographic factors, and the critical need to understand and remedy these disparities. Our patients were all receiving care at a specialized academic epilepsy center, so the outcome disparities we observed were not due to unequal access to neurologic care *per se*. This suggests that structural barriers and disparities in access to healthcare resources or care delivery persist, and impact health outcomes, even in patients with subspecialist access.

In our multivariable analysis, disparities according to sex, insurance, and income persisted after controlling for other demographic factors, while race was no longer significantly associated with the likelihood of seizure freedom. Race and socioeconomic status are strongly correlated in US society due to past and present structural racism, and this correlation means they may not be independent predictors of health outcomes. In our study, socioeconomic factors (income and insurance) were the stronger predictors of seizure freedom. Notably, the multivariable model also accounted for the number of prescribed medications and the time between clinic visits, meaning that two patients seen at similar intervals, prescribed the same number of antiseizure medications, have different seizure freedom likelihoods based in part on socioeconomic factors.

Our study had several limitations. Our data do not reveal the cause of the disparities we observed. Future studies must seek to understand the causes and, more importantly, attempt interventions to reduce the disparities that are now widely recognized throughout healthcare. Our demographic categories were limited by the variables in our electronic health record, which used outdated frameworks for race and gender, and did not capture the full diversity of our patient population. Our study was performed at a US academic medical center with a presumed bias towards more difficult and complex epilepsies. A possible limitation of our analysis of NLP model bias is that, if biases were present in the human annotations used for both training and testing our NLP model, this could be internalized by the model and not detected in its performance on the testing set. However, our human annotations were performed in triplicate by independent readers who were unaware of the identities or demographics of the patients, so bias at that step is unlikely.

## Conclusions

In conclusion, seizure outcomes extracted by natural language processing revealed disparities in the likelihood of seizure freedom across many social determinants of health. We found no evidence of intrinsic model biases within the NLP algorithm. These findings highlight the critical need to reduce disparities across healthcare.

## Acknowledgements

This research was funded by the National Institute of Neurological Disorders and Stroke DP1NS122038; by the National Institutes of Health R01NS125137; the Mirowski Family Foundation; by contributions from Neil and Barbara Smit; and by contributions from Jonathan and Bonnie Rothberg. WKSO was supported by the National Science Foundation Research Grant Fellowship DGE-1845298. RSG was supported by the National Institute of Neurological Disorders and Stroke T32NS091006. CAE was supported by the National Institute of Neurological Disorders and Stroke of the National Institutes of Health Award Number K23NS121520; by the American Academy of Neurology Susan S. Spencer Clinical Research Training Scholarship; and by the Mirowski Family Foundation. DR’s work was partially funded by the Office of Naval Research Contract N00014-19-1-2620.

## Conflict of Interest and Ethical Publication Statement

Authors have no competing interests to disclose. We confirm that we have read the Journal’s position on issues involved in ethical publication and aℒrm that this report is consistent with those guidelines.

## Author Contributions

KX, WKSO, RSG, DR, BL, CAE conceptualized and implemented the study. KX, WKSO, RSG, and CAE had access to and have verified the data reported in the manuscript. AL, CEH, RHH, and KBJ provided feedback on the methods, design, and manuscript of the study. All authors were involved in the drafting, editing, and approval of the final manuscript.

## Data Availability Statement

Our NLP models are available on the Hugging Face hub at https://huggingface.co/CNT-UPenn, and our code is available on GitHub at https://github.com/penn-cnt/NLP_Disparities_in_Seizure_Freedom. We do not make our data available to protect patient privacy.

