## Supplemental Information for "Disparities in seizure outcomes revealed by large language models"

### Supplemental material for: Disparities in Seizure Outcomes Revealed by Large Language Models

### Methods

*Algorithmic Outputs*

Our NLP algorithm was designed for trinary classification: if information regarding the patient’s seizure freedom was contained in the note, it would classify that patient at that visit as either seizure free, or having seizures. If that information did not exist in the note, then the algorithm would give a third category for insufficient information. In our univariable and multivariable analysis of disparities in seizure outcomes - as well as in our initial bias assessment - we obviated notes with a classification of insufficient information.

The confidence of a prediction is an important consideration when applying a model in a clinical setting. For each sample we applied a logit function to the softmax outputs to extract the predicted probability of each of the three classes: seizure free, had seizure, or insufficient information.

### Results

*Visualizing our patient cohort*

We show how many patients we see from each zip code normalized by the number of people per zip code (supplemental figure 1); the proportion of non-male patients from each zip code; the binary (white vs. non-white) racial composition of patients from each zip code (supplemental figure 2); the mean age of our patients from each zip code (supplemental figure 3); and the average seizure freedom rate across visits from each zip code (supplemental figure 4). For reference, we also map the median household income for each zip code, the binary racial composition per zip code, and the number of persons per zipcode using data taken in the 2020 census (supplemental figures 5).

### Supplementary Table 1: Model bias analyses.

| **Variable** | **Metric** | **P-Value** | **Adjusted P-Value** |
| --- | --- | --- | --- |
| **Sex** | Accuracy | 0·64 | 0·96 |
|  | PCB | 0·34 | 0·86 |
|  | NCB | 0·63 | 0·96 |
| **Race** | Accuracy | 0·91 | 0·96 |
|  | PCB | 0·91 | 0·96 |
|  | NCB | 0·43 | 0·86 |
| **Ethnicity** | Accuracy | 1·00 | 1·0 |
|  | PCB | 0·81 | 0·96 |
|  | NCB | 0·39 | 0·86 |
| **Insurance** | Accuracy | 0·16 | 0·86 |
|  | PCB | 0·34 | 0·86 |
|  | NCB | 0·15 | 0·86 |
| **Median Zip-Code Income** | Accuracy | 0·37 | 0·86 |
|  | PCB | 0·70 | 0·96 |
|  | NCB | 0·24 | 0·86 |
| **Age** | Accuracy | 0·31 | 0·86 |
|  | PCB | 0·74 | 0·96 |
|  | NCB | 0·78 | 0·96 |
